# Safety, tolerability and efficacy outcomes of the Investigation of Levetiracetam in Alzheimer’s Disease (ILiAD) trial

**DOI:** 10.1101/2024.05.14.24307228

**Authors:** Arjune Sen, Sofia Toniolo, Xin You Tai, Mary Akinola, Mkael Symmonds, Sergio Mura, Joanne Galloway, Angela Hallam, Jane Y C Chan, Ivan Koychev, Chris Butler, John Geddes, Gabriel Davis Jones, Younes Tabi, Raquel Maio, Eleni Frangou, Sharon Love, Sian Thompson, Rohan Van Der Putt, Sanjay G Manohar, Rupert McShane, Masud Husain

## Abstract

**Objective:** To assess whether the antiseizure medication levetiracetam may improve cognition in individuals with Alzheimer’s disease who have not previously experienced a seizure.

**Methods:** We performed a randomised, double-blind, placebo-controlled crossover study in individuals with mild-to-moderate Alzheimer’s disease. Electroencephalography was performed at baseline and those with active epileptiform discharges were excluded. Eligible participants were randomised to placebo for 12 weeks or an active arm of oral levetiracetam (4 weeks up-titration to levetiracetam 500 mg twice daily, 4 weeks maintained on this dose followed by 4 weeks down-titration to nil). Participants then crossed over to the other arm. The primary outcome was change in cognitive function assessed by the Oxford Memory Task, a task sensitive to hippocampal memory binding. Secondary outcomes included tolerability, other neuropsychological scales and general questionnaires.

**Results:** Recruitment numbers were severely limited owing to restrictions from the COVID-19 pandemic at the time of the study. Eight participants completed both arms of the study (mean age 68.4 years [SD=9.2]; 5 females [62.5%]).

No participants withdrew from the study and there was no significant difference between reported side-effects in the active levetiracetam or placebo arm. Measures of mood and quality of life were also not significantly different between the two arms based on participant or carer reports. In limited data analysis, there was no statistically significant difference between participants in the active levetiracetam and placebo arm on the memory task.

**Significance:** This pilot study demonstrates that levetiracetam was well tolerated in individuals with Alzheimer’s disease, who do not have a history of seizures, and has no detrimental effect on mood or quality of life. Larger studies are needed to assess whether levetiracetam may have a positive effect on cognitive function in subsets of individuals with Alzheimer’s disease.

**Plain language summary:** Abnormal electrical activity within the brain, such as is seen in seizures, might contribute to memory problems in people with dementia. We completed a clinical trial to see if the anti-seizure medication, levetiracetam, could help with memory difficulties in people with Alzheimer’s disease (the most common cause of dementia). In this pilot study, we could not prove whether levetiracetam helped memory function. We did show that the drug is safe and well-tolerated in people with dementia who have not had a seizure. This work therefore offers a platform for future research exploring anti-seizure medications in people with dementia.

**Key points:**

1. Epilepsy may contribute to the aetiopathogenesis of Alzheimer’s disease
2. ILiAD is a double-blind, randomised placebo controlled trial of levetiracetam in Alzheimer’s disease
3. Levetiracetam was well tolerated in people with AD who have not had a seizure
4. Owing to small sample size, effect of levetiracetam on cognition could not be determined
5. Larger trials of anti-seizure medications in people with dementia are warranted

## 1. Introduction

The burgeoning healthcare needs of dementia threaten to overwhelm health care systems around the world. In the United States, for example, there are estimated to be over 6 million people living with dementia leading to a current cost of over 345 billion dollars.**^1^** Over the next 40 years it is predicted that there will be around 13 million people with Alzheimer’s disease (AD) in the United States alone with most of this increase occurring in people aged over 85 years. Dementia prevalence is also increasing in low to middle income countries where populations are ageing three times faster than in the global west.**^2^** In such resource-poor settings the direct and indirect impacts of dementia are vast and it will be very difficult for people living in such settings to access highly specialised and expensive medicines that may only offer modest improvements in cognitive function.

Epilepsy also increases as populations age with its greatest incidence in those over 75 years of age. Although it has been recognised for many decades that seizures are more common in people with dementia,**^3^** until recently this has been considered an epiphenomenon – a simple consequence of neuronal loss destabilising neuronal networks. Over the past decade, several lines of enquiry have demonstrated that there is a bidirectional relationship between epilepsy and dementia.**^4–6^** It seems likely that seizures or epileptic activity in the ageing brain may contribute to the pathogenesis of AD.**^7–10^** Importantly, epilepsy can often be treated with cheap, generic antiseizure medications (ASMs).**^11^** The inter-linking of epilepsy and dementia therefore raises the possibility that suppression of abnormal epileptic activity may help cognition in those with AD.

ASMs have been explored in animal models of AD with a particular focus on the synaptic vesicle 2A antagonist levetiracetam.**^12^** Levetiracetam can reduce neuritic plaque formation,**^13^** and suppress abnormal electrical activity in animal models of AD. In transgenic mice, levetiracetam also associates with amelioration of cognitive and behavioural deficits. In humans with mild cognitive impairment, suppression of aberrant hippocampal hyperactivation resulted in better performance on a visual recognition task.**^14^** Also, levetiracetam is a widely utilised ASM with minimal drug-drug interactions and demonstrated efficacy and tolerability in older people.**^15^** It, therefore, would seem worthwhile considering whether levetiracetam offers benefit to people with dementia who have *not* previously experienced a seizure.

The Investigation of Levetiracetam in Alzheimer’s Disease (ILiAD) trial was designed as a randomised, double-blind, placebo-controlled crossover study. The primary endpoint was change in cognitive function assessed through a highly specific hippocampal binding task. Secondary endpoints included adverse effects, impact on mood, quality of life assessments and evaluation of electroencephalographic recording to try and predict drug response. Regrettably, the COVID-19 pandemic affected the study very adversely and the trial, which by definition was in a vulnerable older population, had to be halted early. Here we present the data of all participants who completed the study with specific emphasis on how similar studies can be developed in future.

## 2. Methodology

### 2.1 Main trial outline

The methodology underpinning the ILiAD (registration number: NCT03489044) trial has been published previously.**^16^** In brief, people aged over 50 years with a confirmed diagnosis of mild to moderate AD, who had not experienced an overt seizure, were invited to participate.

Participants were recruited from Oxford University Hospitals NHS Foundation Trust and Oxford Health Foundation Trust. The clinical team made initial approaches. All participants had to have capacity to provide informed consent and had to have a named carer who would assist with trial procedures.

Potentially suitable candidates underwent initial screening that included checks of renal function, baseline cognitive assessment and an electroencephalogram (EEG). Full inclusion and exclusion criteria are listed in Supplementary Table 1. People with epileptiform abnormalities on EEG recording were specifically excluded as was anybody who was taking an ASM for any reason (for example gabapentin being taken for pain). These strict criteria were applied so that levetiracetam was not simply treating seizure activity and any potential impact on neural networks was not masked/modulated by other ASMs.

Consenting participants who met all inclusion and exclusion criteria were enrolled into the study and randomised 1:1 to begin with levetiracetam or placebo. Levetiracetam (250mg tablets) and placebo were manufactured to look identical and carers dispensed tablets according to a specifically-created treatment chart.**^16^** For those allocated to receive active drug first, levetiracetam was up-titrated by 250mg (one tablet) every week for 4 weeks before being maintained at levetiracetam 500mg (two tablets) twice daily for four weeks. Levetiracetam was then down-titrated to nil over four weeks. People subsequently crossed over to the placebo arm where a similar titration schedule was completed. For those initially allocated to the placebo arm the titration schedule was identical except that placebo was up-titrated and weaned first, followed by the cross over to active drug.

The trial flowsheet (**Figure 1**) was identical for all participants. Key assessment points were at baseline, 8 weeks (after completion of four weeks on active drug/placebo in arm 1) and 20 weeks (after completion of four weeks on active drug/placebo in arm 2). Intra-participant measures were evaluated and group analyses performed.

**Figure 1.**
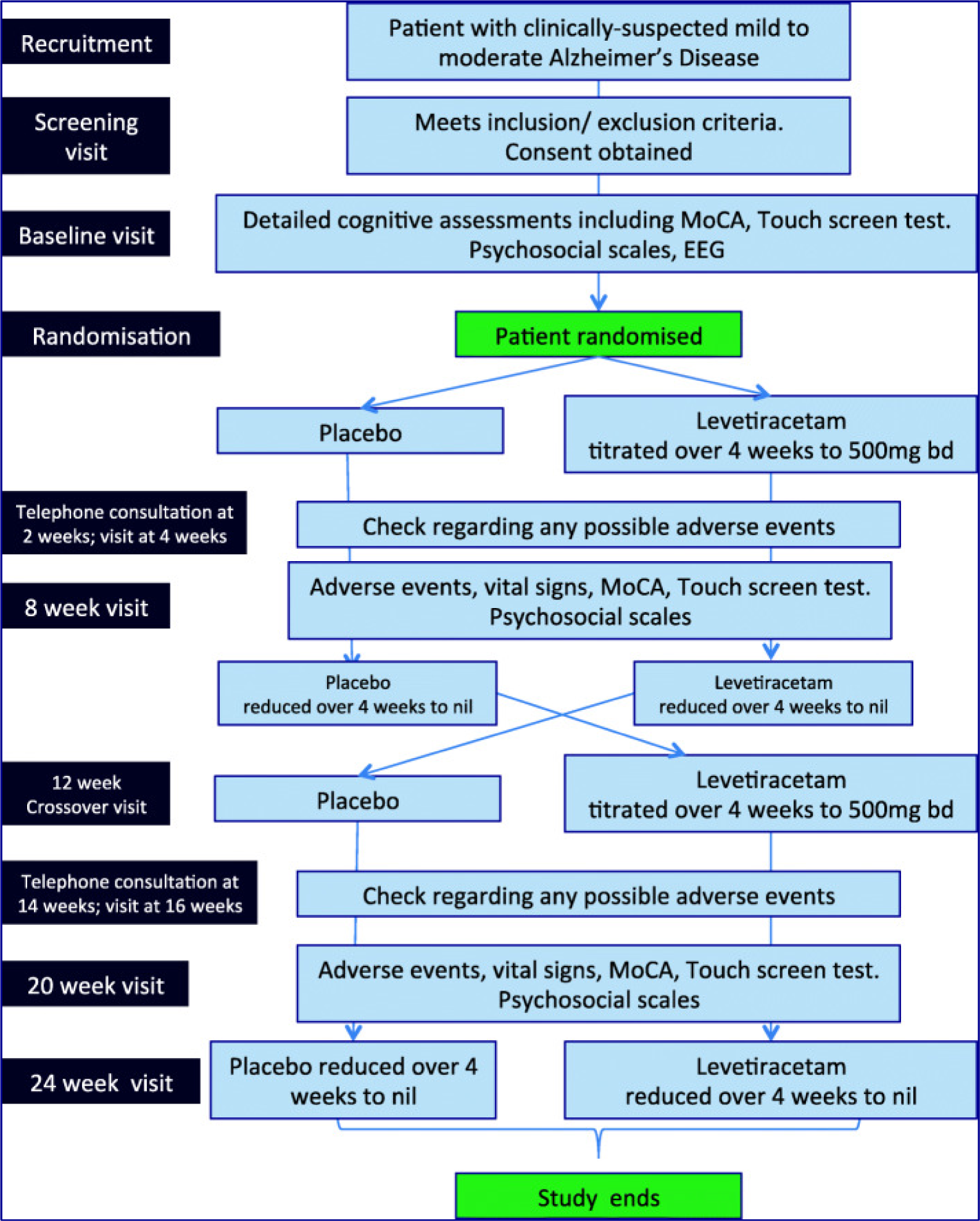
Schema for the ILiAD trial. ILiAD trial flowsheet from recruitment through cross-over and monitoring as published previously.^16^ After baseline assessments, recruited participants were randomised to first receive either levetiracetam or placebo. Detailed assessments were performed at 8 weeks to compare to baseline data. After weaning away levetiracetam/placebo, participants crossed over to the second arm and received the opposite of what they had been given in the first arm (namely people receiving levetiracetam now received placebo and vice versa). Assessment at 20 weeks was compared to data acquired at 12 weeks and levetiracetam/placebo was then down-titrated to nil by 24 weeks.

### 2.2 “What was where?” Oxford Memory Task – primary outcome measure

The ILiAD study used a modified version of the “What was where?” Oxford Memory Task (OMT).**^17,18,19,20,21,22,23^** This task is sensitive to early signs of working memory deficits in individuals with hippocampal dysfunction such as patients with limbic encephalitis,^17^ previous temporal lobe lobectomy,^23^ familial Alzheimer’s AD,^18,22^ late-onset sporadic AD^19,21^ and subjects at risk of developing AD.^19,20^ The OMT can detect early impairment in memory binding even when overall performance is still intact.^17,23^

During the OMT, participants were presented with either one (*Fractals 1*) or two (*Fractals 2*) fractals located randomly on the black screen (**Figure 2**). Participants were asked to remember the design of the fractal(s) shown (‘what’), and their locations (‘where’). After a four second blank screen delay, two fractals appeared at the center of the screen along the vertical midline. One of which was present in the previous array (target) while the other fractal was new (distractor). Participants were asked to touch the target fractal and drag it to the original location. Trial order was randomized within each block and fractal location was pseudorandomized, based on a Matlab script (MathWorks, Inc).

**Figure 2.**
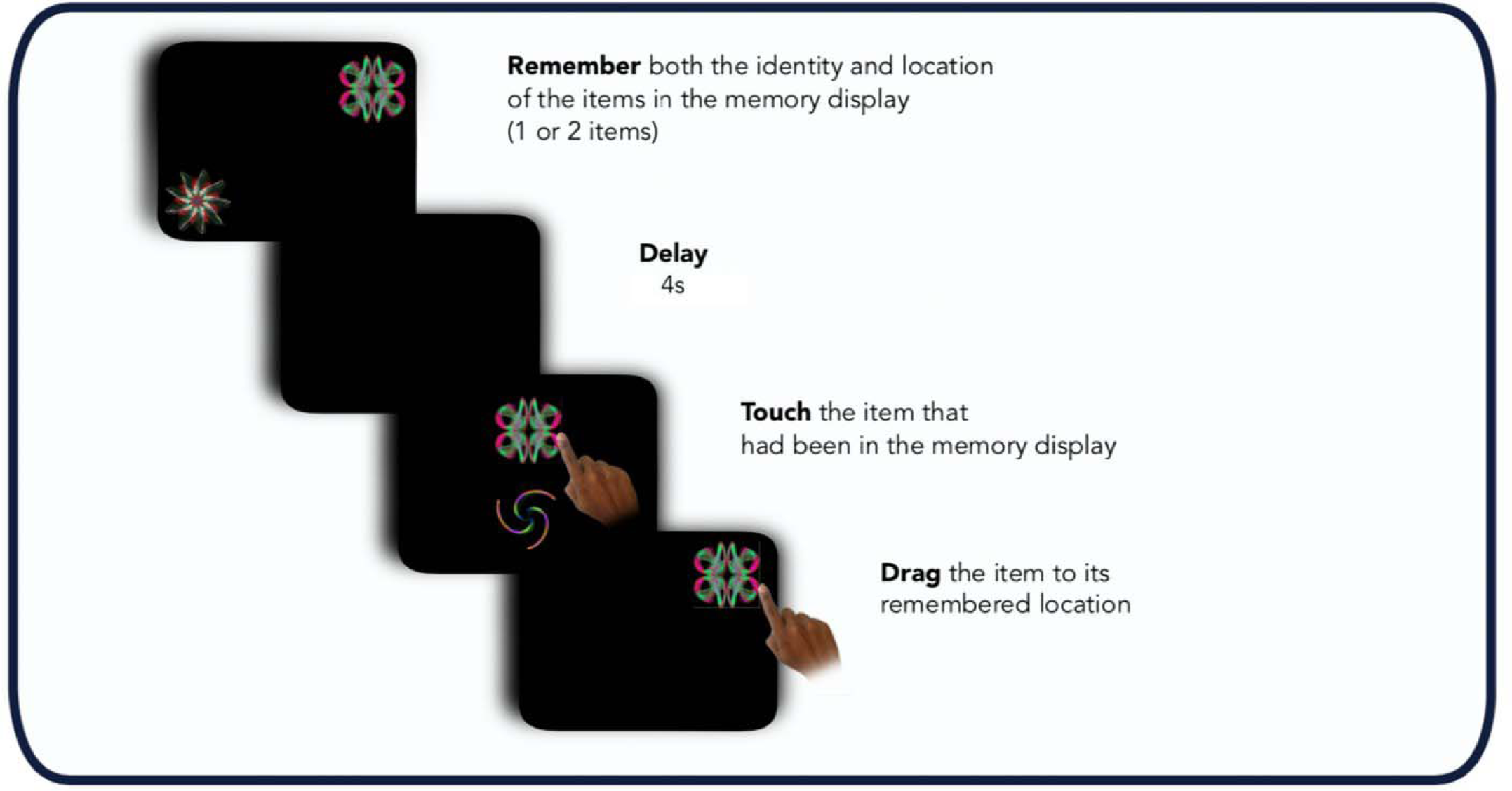
“What was where?” Oxford Memory task (OMT) Participants were presented with either 1 or 2 fractals randomly distributed on the screen. After a 4 second delay two fractals appeared at the center of the screen, one of which had appeared in the memory array whereas the other one was a distractor. Firstly, they needed to identify the object they had seen previously (‘what’), and then drag it back to its original location (‘where’).

Stimuli were presented on a black background and were chosen from a library of 196 fractals (http://sprott.physics.wisc.edu/fractals.htm). Participants sat approximately 30 cm in front of a tablet (either iPad or Android), yielding 2.3° of visual angle. Stimuli were calibrated using the dimension on the screen to ensure matching of stimuli properties across different tablet models.

Several primary working memory metrics can be determined from the OMT (**Figure 2**), including:

- **Identification Time**: the time in seconds taken to identify the correct object.
- **Localization Time** : the time in seconds to drag the chosen object to its remembered location.
- **Proportion correct** : the proportion of trials in which participants correctly identified the target.
- **Absolute Error** : the distance from the center of original item location to the center of participant’s response location.

In the two-item condition (Fractals 2), further metrics can be derived based on the Mixture Model of working memory by Bays et al ^24^, adapting the original model by fitting the data to different memory outcome distributions using a permutation approach, described further in **Supplementary** Figure 1. This approach has been previously published using this task.**^25,26^**

- **Target detection:** the probability of correctly identifying the target.
- **Guessing**: the probability of random guessing responses.
- **Misbinding**: the probability of mislocalizing correctly identified item to the remembered location of another item in the memory array.
- **Imprecision**: the width of the distribution of the responses around the target.

Based on power calculations (90% power at a 5% significance level),**^16^** a total of 24 participants would be needed to detect a meaningful difference on the OMT. To account for an attrition rate of up to 20%, we planned to recruit 30 participants.

### 2.3 Neuropsychological tests, questionnaires and scales

The tests and questionnaires administered during ILiAD included the Mini Mental State Examination (MMSE)^27^, Bristol Activity of Daily Living Scale (BADLS)^28^, Dementia Severity Rating Scale (DSRS)^29^, Neuropsychiatric Inventory (NPI)^30^, Euro-Qol Quality of Life measure (EQ5D)^31^ and Quality of life scale (QoL)^32^. QoL scales included a patient version (patient responded regarding his/her quality of life) and a caregiver version (caregiver responding on their quality of life). EQ5D scales included a patient version, a caregiver version and a proxy version (caregiver responding on the patient’s estimated quality of life).

### 2.4 Electroencephalography recording

A baseline EEG was performed for all potential participants, principally to exclude the presence of epileptiform activity. Recordings were obtained via scalp electrodes, individually attached according to the international 10-20 system, following the local hospital protocols.

### 2.5 Adverse effect reporting

Levetiracetam, as outlined, is a well-tolerated and widely prescribed anti-seizure medication and is a drug of choice in older people with epilepsy. The safety and tolerability profile of levetiracetam is well-established. A full description of our adverse event reporting and harms protocol has been provided previously with principal potential adverse events listed in **Supplementary table 2**.**^16^**

### 2.6 Statistical analyses

As outlined in the ILiAD research protocol^16^ (**Figure 1**), changes in questionnaire responses were calculated by comparing the change from the end of each arm of the study to its baseline, i.e. week 8 compared to baseline (arm 1) and week 20 compared to week 12 (arm 2). For demographics and baseline cognitive tests (MMSE), a two-sample t-test or Mann-Whitney U test were used to compare continuous variables, according to the normality of the data. Chi-square test was used to compare categorical variables. Paired t-test and Wilcoxon signed-rank test were used to compare questionnaires’ measures between the two arms of the study, according to the normality of the data.

Given the final small sample size, which was not anticipated when conceiving the study, for the computerized metrics we fitted a within-subject linear mixed effect model, as is standard practice in experimental psychology for experiments with small number of participants studied under two or more experimental conditions.**^33^** The interindividual variability was taken into account by including trial by trial data (20 trials per session) for each individual and by considering each participant uniquely in the model, whilst looking at the separate effects of session (by comparing the first and the second session) and the effect of drug (placebo or levetiracetam) across subjects. For each outcome variable, the following model was fitted: var ∼ 1 + session*drug + (1 | subject). Significance was set at p-value < 0.05.

## 3. Results

### 3.1 Randomisation

Prior to having to halt the trial, 8 participants were recruited. No participant was unblinded for clinical purposes before the end of the trial. All participants completed the entire study – both arms. Owing to the pandemic, though, the trial had to be adapted and certain tasks that required in-person assessments could not be completed.

At the end of the study, unblinding revealed that 3 individuals were allocated to the arm where placebo was given first, and 5 to the arm where levetiracetam was given first (**Table 1**).

**Table 1.** Demographics overview. Values are presented as mean, with standard deviation (SD) in brackets. M = male, F = female. MMSE = Mini mental state examination. LEV = Levetiracetam. n.s. = not significant. OMT = “What was where?” Oxford Memory task. The different OMT timings reflect the four different timepoints where OMT was scheduled in the trial, with the relative number of subjects completing the OMT test at each timepoint.

|  | Overall<br>(n = 8) | PLACEBO first<br>(n= 3) | LEV first<br>(n = 5) | p-value |
| --- | --- | --- | --- | --- |
| Age | 68.4 (9.2) | 70.3 (6.0) | 67.2 (11.2) | n.s. |
| Sex (M/F) | 3/5 | 0/3 | 3/2 | n.s. |
| MMSE | 17.7 (4.5) | 15.3 (3.2) | 19.2 (4.8) | n.s. |
| Dropout/withdrawn | 0/8 | 0/3 | 0/5 |  |
| Questionnaires – 20 weeks | 8/8 | 3/3 | 5/5 |  |
| OMT - Baseline | 8/8 | 3/3 | 5/5 |  |
| OMT - 8 weeks | 4/8 | 2/3 | 2/5 |  |
| OMT - 12 weeks | 3/8 | 2/3 | 1/5 |  |
| OMT - 20 weeks | 0/8 | 0/3 | 0/5 |  |

### 3.2 Demographics

There was no difference between participants in the two arms (people started on levetiracetam first vs. placebo first) in terms of age, sex, or baseline MMSE (**Table 1**). As the trial was interrupted owing to the COVID-19 pandemic and given the necessary face-to-face nature of the MMSE test, this was administered only at baseline. All participants completed the other questionnaires and scales remotely until the end of the study (week 24).

The “What was where?” OMT also required in person assessment. For the OMT, data collection was interrupted at different stages of enrolment (**Table 1)**. Datapoints of the OMT task from week 12 onwards were excluded from further analyses as none of the participants completed the task at week 20. Data from those with only OMT assessment at baseline were also excluded from further analyses. As a result, the samples for assessment of the primary endpoint compared where n = 2 per arm with 2 timepoints per arm.

### 3.3 Primary outcome: “What was where?” Oxford Memory task (OMT)

Overall, there was no statically significant difference in the metrics examined between participants in the two arms (started on levetiracetam first or placebo first) for Fractals 1 (**Figure 3**, **Table 2**), and Fractals 2 (**Supplementary** Figures 2 and 3**, Supplementary Table 3**). The very small numbers, though, do not allow firm conclusions on this.

**Figure 3.**
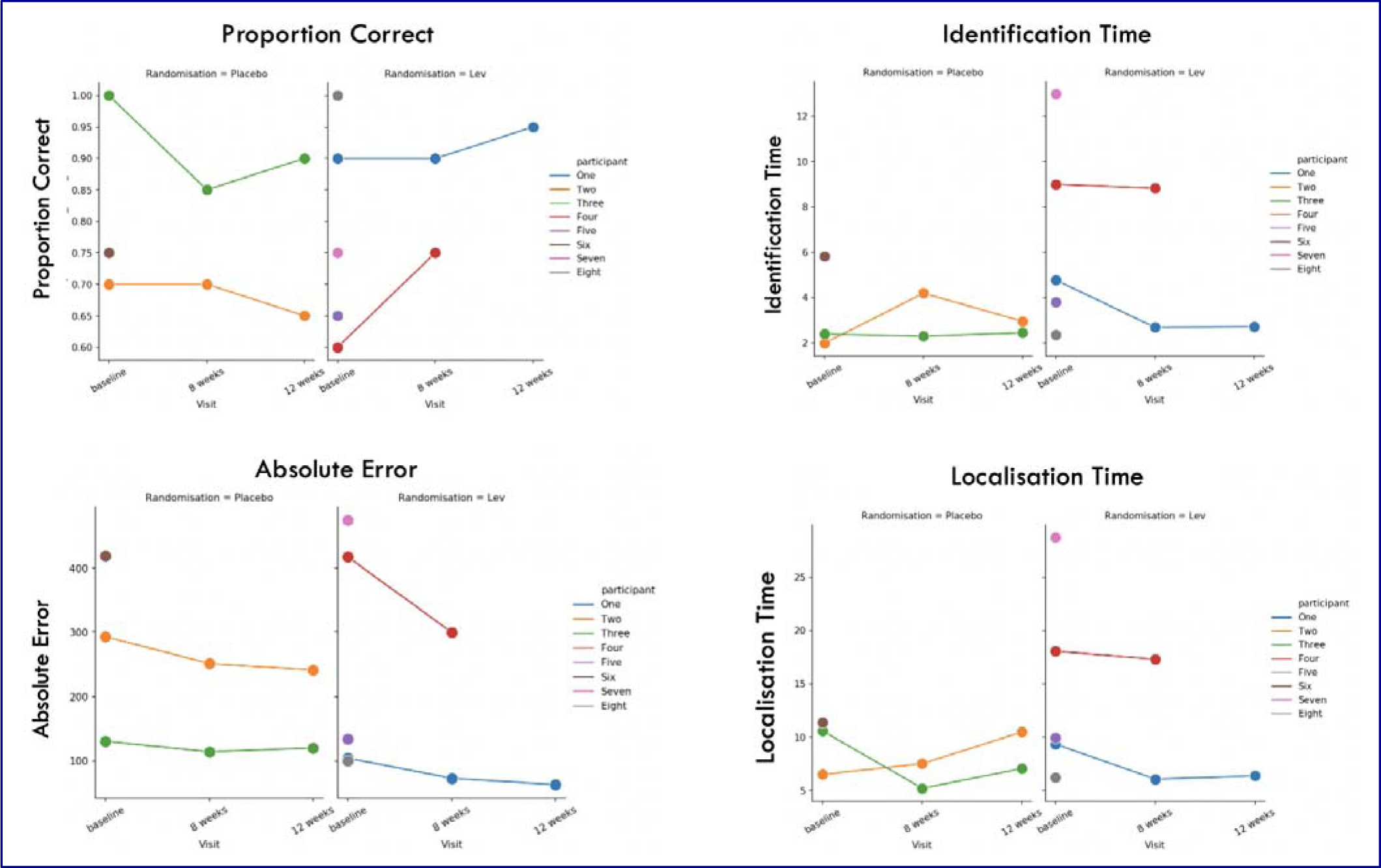
Individual datapoints for Fractals 1 testing. Values are presented as color-coded individual datapoints, for all subjects who took part in the study. Only data from patients who completed at least one arm of the study were retained, and datapoints for only one arm of the study were retained per subject as none completed 4 sessions 9 (both arms). Randomization status is represented by different subplots for each metric (left for placebo, right for levetiracetam). Lev = levetiracetam. Proportion correct = proportion of correctly identified items. Identification time = time in seconds identify the correct object. Absolute Error = distance between the original item location to the participant’s response location. Localisation Time = the time in seconds to drag the chosen object to its remembered location. More positive values correspond to better performance for proportion correct, and to worse performance for absolute error, identification and localisation time.

**Table 2.** Fractals 1 Results. Session = baseline or 8 weeks, Drug = randomization allocation (whether on placebo or levetiracetam), Session × Drug = interaction between session and drug, i.e., whether being on the levetiracetam or placebo changed the cognitive outcome. P = p-value, with statistical significance set at p < 0.05. The significant results for session * drug interaction for Identification time did not survive multiple comparisons’ correction.

|  | Coefficients | Standard Error | t-value | p-value |
| --- | --- | --- | --- | --- |
| **Fractals 1** |  |  |  |  |
| **Proportion correct** |  |  |  |  |
| Session | -0.07 | 0.089 | -0.84 | 0.40 |
| Drug | -0.25 | 0.20 | -1.23 | 0.21 |
| Session * Drug | 0.15 | 0.13 | 1.20 | 0.24 |
| **Absolute Error** |  |  |  |  |
| Session | -22.46 | 34.13 | -0.66 | 0.51 |
| Drug | 97.55 | 116.6 | 0.84 | 0.40 |
| Session * Drug | -60.70 | 48.3 | -1.26 | 0.21 |
| **Identification time** |  |  |  |  |
| Session | -505.53 | 2251.2 | -0.22 | 0.82 |
| Drug | 10033 | 3183.7 | 3.15 | 0.002 |
| Session * Drug | -4696.3 | 2013.5 | -2.33 | 0.02 |

### 3.4 Secondary outcomes

#### 3.4.1 Safety and tolerability outcomes Study dropouts and missing doses

No participants withdrew from the study. One participant delayed increasing from one tablet to two tablets by three days due to a participant query. After a telephone consultation, it was deemed suitable to continue the participant in the study, and the dose was increased up to the target dose. No other missing doses neither reported at any time by any other participants nor revealed by the drug administration chart.

#### 3.4.2 Neuropsychological scales and questionnaires

#### Measures of mood, activity of daily living, dementia severity, and quality of life

There was no statically significant difference between placebo and levetiracetam in measures of mood (NPI), activity of daily living (BADLS), dementia severity (DSRS), and quality of life (QoL, EQ-5D, DSRS), whether reported by the patient, by the caregiver, or if estimated by the caregiver with respect to the patient (**Supplementary** Figure 4).

#### 3.4.3 EEG recordings

Baseline EEG recordings were assessed by a consultant neurophysiologist (MS) for any clinically relevant findings. There was no evidence of epileptiform activity in any study participants. Planned further analyses of baseline EEG to predict response was not performed owing to the small numbers recruited.

#### 3.4.4 Adverse events reported

There were 35 adverse instance reports that ranged from “fatigue” (three instances) to “word finding difficulty” (two instances). “Headache” was the most frequently reported symptom with 11 instances (**Supplementary** Figure 5). There were no suspected unexpected serious adverse reactions (SUSARs) reported.

Three of the eight study participants accounted for 33 out of 35 adverse instance reports. Of these reports, 19 were made whilst the participants were taking placebo and 16 were reported during whilst the participant was on levetiracetam (**Figure 4**). There was no difference in number of adverse reporting instances between active and non-active arms of the study (t= -0.243, 95%CI - 8.32 – 6.82, p=0.82)

**Figure 4.**
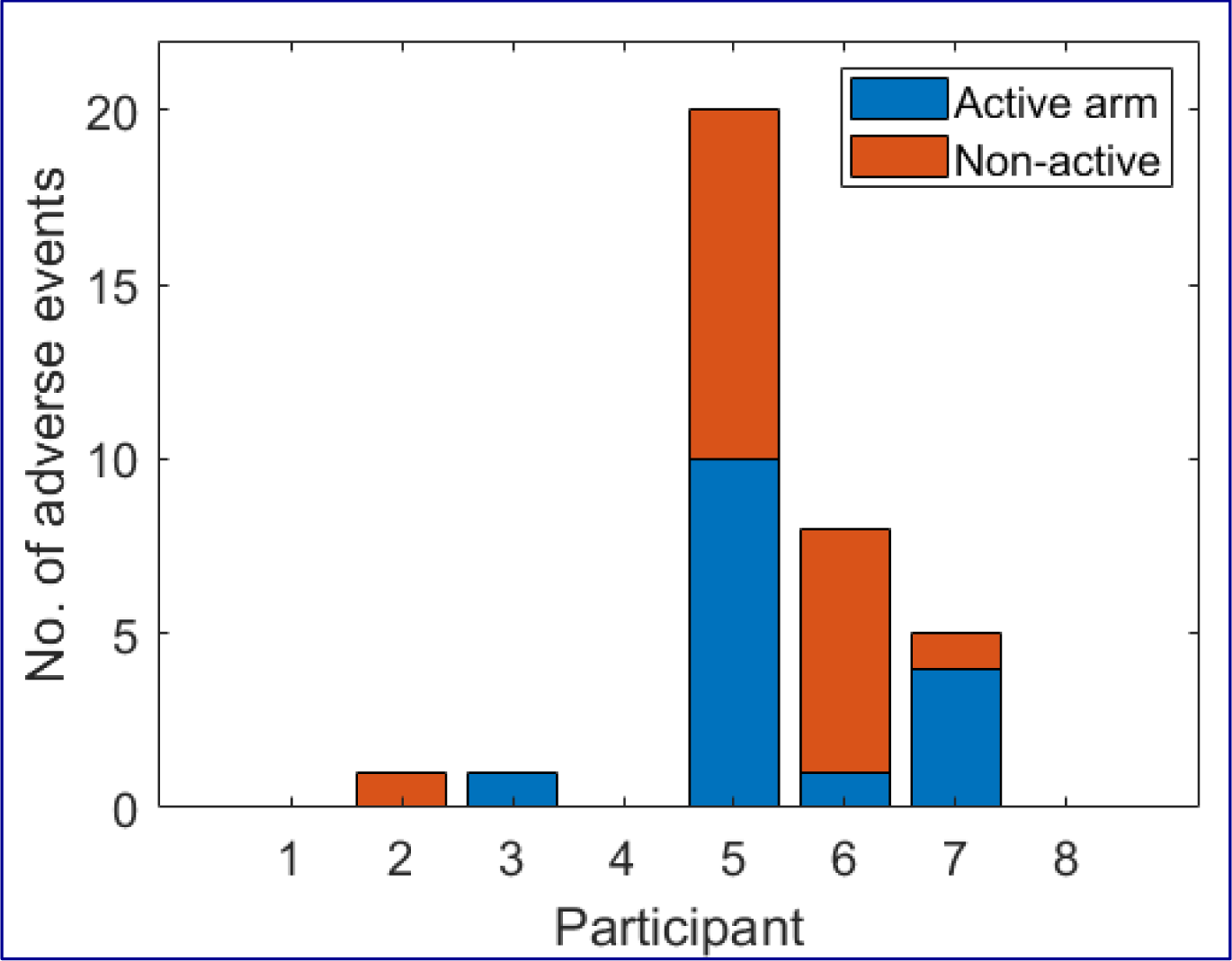
Number of adverse reporting instances. Four study participants accounted for all the adverse reporting instances in the study (total 5). Nineteen adverse instances were reported whilst participants were in the non-active, placebo arm (red) of the study and 16 were reported during the active, Levetiracetam arm (blue).

## 4. Discussion

In this proof-of-concept study we assessed the safety and tolerability of levetiracetam in patients with AD without overt seizures or baseline epileptic EEG activity, and the effect of levetiracetam on cognition. Regrettably, we were not able to complete recruitment owing to the COVID-19 pandemic.

The main result of this trial is that levetiracetam is well tolerated in this patient population, namely older people with AD who have not experienced a seizure. No participant withdrew from the study prematurely. The adverse effects were mild, in line with known side effects of levetiracetam^34^ and there was no difference in the rate of side effects when comparing levetiracetam to placebo. Importantly, there was no worsening of mood in the levetiracetam arm compared to the placebo arm, both in terms of self-reported adverse effects and objective measures such as the NPI. This reinforces previous data showing levetiracetam as a safe and well tolerated drug in older people ^34-38^, and extends these findings specifically to individuals with AD.

We did not have sufficient data to assess the primary outcome of the study, the effect of levetiracetam on cognition, measured by our computerized cognitive task. All participants were, though, able to complete the in-person computerized task when it was possible to run the test, with no incomplete data. As those recruited were primarily in a moderate stage of the disease, as indexed by the MMSE (**Table 1**), our data suggests that this computerized task can be a viable resource to quantify granular changes in cognition over a short time scale. Such tests measure working memory and are less susceptible to practice effects compared to traditional tests of cognition. Future trials in AD might consider similar cognitive testing strategies to substantially reduce trial length.

Titration of IMP in ILiAD was relatively brisk and levetiracetam was increased to 500mg twice daily, higher than was employed in previous studies of MCI and a similar study in AD.**^14,39^** Our initial rationale was that people with AD are likely to have more network instability than those with MCI, hence a higher dose of levetiracetam might be required in AD. While we cannot comment on the effect of this higher dose on the primary outcome measure, it is serendipitous that we can show that there are no adverse neuropsychiatric effects from levetiracetam 500mg twice daily compared to placebo.

A similar, independent study was run concurrently with the ILiAD trial to address whether levetiracetam could improve executive function in patients with Alzheimer’s disease **^39^**. In that American cohort, 34 individuals with Alzheimer’s disease completed a similar double-blind, cross-over study design with a substantially lower dose of levetiracetam (125 mg twice daily). That study’s primary endpoint was not met as no overall difference in cognitive outcomes were detected with levetiracetam compared to controls. Nine patients, however, who had detectable epileptiform activity on their baseline EEG, showed a small improvement on a Stroop test subscale and virtual route learning test at the group level suggesting a potential domain-specific improvement in executive skills **^38^**. Consistent with the ILiAD trial, there were no safety concerns reported **^39^**.

As outlined, participants with epileptiform activity were excluded from ILiAD. Suppression of seizures is the first step to helping cognitive function in those with epilepsy. Given that AD so closely associates with seizure disorders, it is possible that people with AD and epileptiform activity on EEG recording have experienced unrecognized seizures and that, therefore, in that sub-population levetiracetam is simply treating a hitherto unrecognized epilepsy. Taken together, though, these two innovative trials offer a new avenue to trying to improve cognitive function in AD and confirm that levetiracetam is a safe medication in this patient cohort.

## 5. Limitations

Limitations of this study are primarily driven by the premature interruption of the trial, which resulted in a small number of individuals being recruited (8 compared to 30 planned) and lack of completion of the computerized cognitive tasks (only 2 subjects per arm and only 2 timepoints per arm). Since the inception of ILiAD we have developed a fully remote version of the “What was where?” Oxford Memory task which is now available at https://oxfordcognition.org/. In this way, our computerized cognitive assessment can now be performed regardless of the nature of the visit (remote or in-person). This new version of the OMT is deployable across devices (computers, tablets and mobile phones), thus resulting in higher scalability of future, possibly multicentric clinical trials.

The small sample number and lack of being able to test the primary endpoint, also meant several subsidiary analyses could not be performed – for example whether analysis of the EEG might provide a biomarker for who could benefit most from levetiracetam therapy.

While disappointing that such hypotheses could not be tested, the effort and reorganisation involved to continue a trial in one of the most vulnerable groups (older people with AD) during the COVID-19 pandemic cannot be underestimated. Many researchers were redeployed to COVID studies or other clinical roles and it is testament to the participants recruited that all individuals completed the study even though there were so many changes to testing and, for example, delivery of IMP.

## 6. Conclusion

These pilot data show that levetiracetam is well tolerated in patients with AD who have not have seizures and has no detrimental effect on mood or quality of life. Larger studies should be very actively explored to assess whether levetiracetam may have a positive effect on cognitive function in those with AD who have not experienced a seizure. Given the rapid increase of people with dementia in low to middle income countries, particular emphasis should be given to trialling levetiracetam in these resource poor settings.

## Data Availability

All data produced in the present study are available upon reasonable request to the authors

## Acknowledgements

This work was supported by a Medical Research Council (UK) Confidence in Concept Grant and by the Investigator Initiated Study Programme of UCB Pharma.

## Declaration

We confirm that we have read the Journal’s position on issues involved in ethical publication and affirm that this report is consistent with those guidelines.

## Data availability statement

Raw data files will be made available on reasonable request

## Ethical Approval

Ethical approval was provided by Oxford Research Ethics Committee B and the Health Research Authority (National Health Service, England; ref no:17/SC/0468).

Clinical trial registration: clinicaltrials.gov - NCT03489044

All participants were required to have capacity to consent to the ILiAD study and such consent was provided by all enrolled participants

## Contributions

AS conceptualised, initiated and was chief investigator for the trial. STo, XYT, MA, MS, SM, JG, IK, STh, RvDP, SM, RMcS, MH were all responsible for recruitment and data acquisition. SM, JG were the clinical trial pharmacists. AH was responsible for dispensing active drug and placebo. STo, XYT, GDJ, YT, RM were involved in analysis of data, this being led by STo and XYT. AS, IK, JC, CB, JG, SM, RMcS, MH provided strategic oversight through the study. AS, STo and XYT drafted the manuscript. Other listed co-authors reviewed and commented on the draft manuscript.

## Preprint

This work will be released as a preprint and is under review at Epilepsia Open

