## Supplementary material for "Safety, tolerability and efficacy outcomes of the Investigation of Levetiracetam in Alzheimer’s Disease (ILiAD) trial"

**The ILiAD Trial: Supplementary materials**

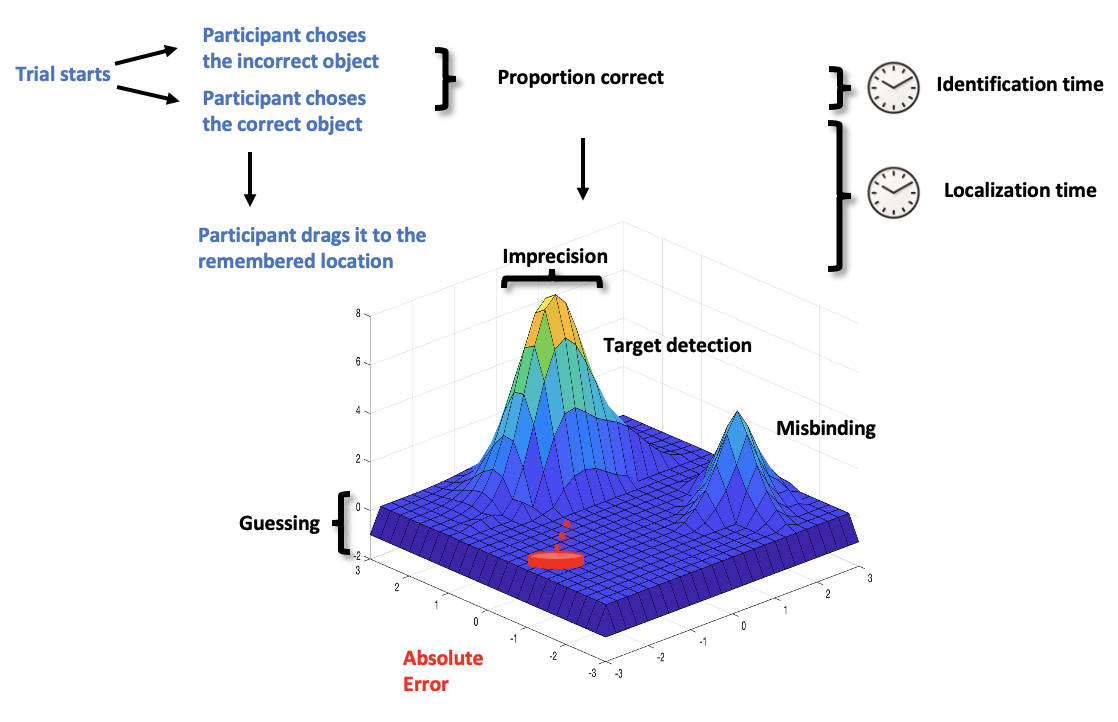

**Supplementary Figure 1 | Summary metrics that can be measured from the “What was where?” Oxford Memory task (OMT)**

Primary metrics of proportion correct, identification time, localization time and absolute error are obtained from the response in each trial. Further metrics are derived in the two-fractal condition by fitting the data to a mixture model to approximate three different distributions to represent guessing, imprecision and misbinding. Model fitting was achieved using a permutation approach, where for each single trial distances between the response location and the location of a target, a distractor (non-target in that trial, i.e., misbinding) and a distractor taken from a randomly chosen trial (i.e., guessing) were calculated. The response was either counted as target (1), distractor (2) or random guessing (3) according to which distance was the shortest. This procedure was repeated 5000 times per trial, introducing a distractor from a randomly chosen trial each time, and the proportions for these three sources of response per trial (absolute amount of response type/5000) were computed. The introduction of a distractor that was randomly chosen from another trial allowed us to differentiate whether an error was systematically linked to the very specific trial’s distractor or whether it could be accounted for even by a randomly chosen distractor that was not present in that specific trial. Importantly, these metrics were calculated uniquely on trials where an object was correctly identified.

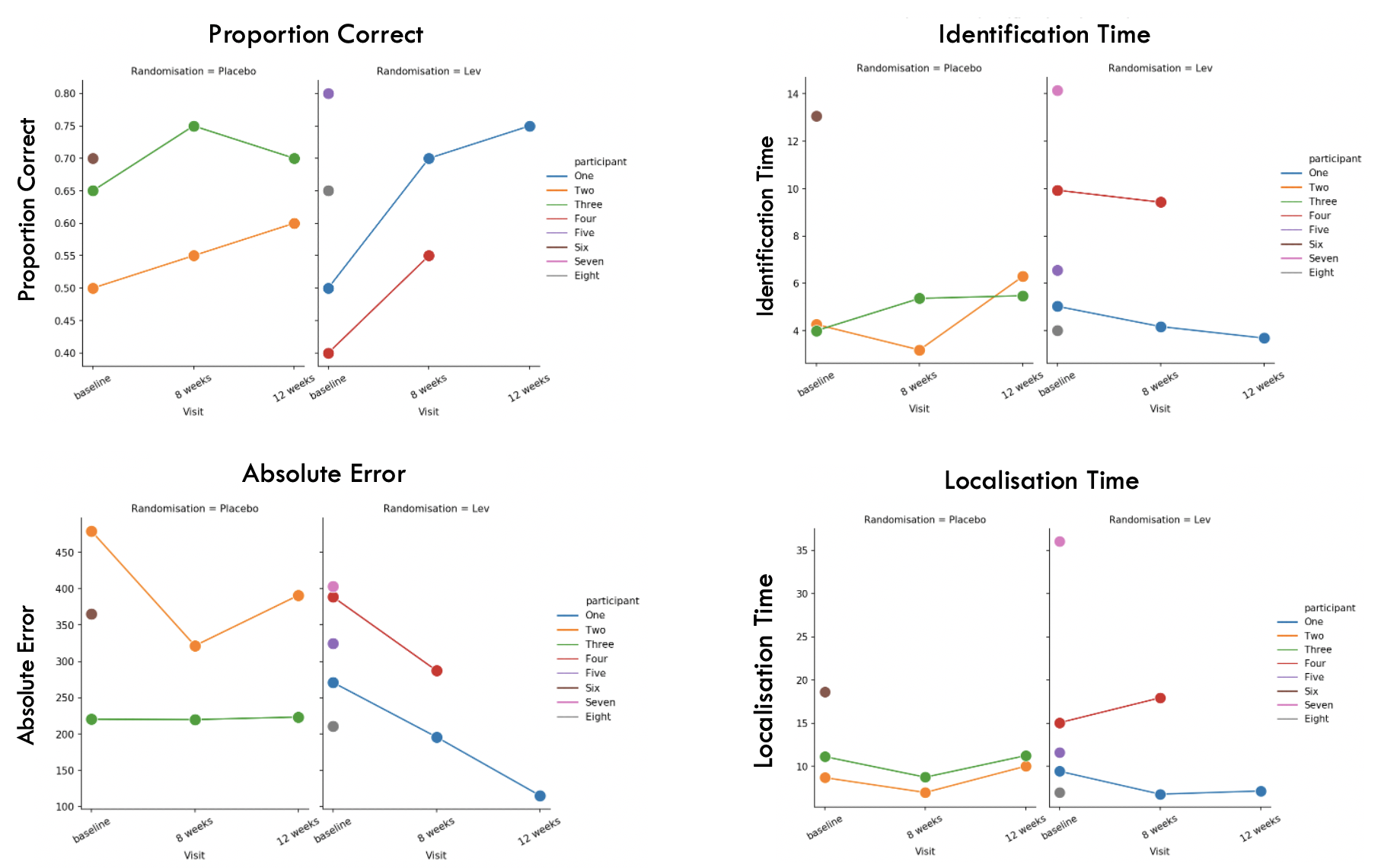

**Supplementary Figure 2| Individual datapoints for Fractals 2 (basic metrics)**

Values are presented as color-coded individual datapoints, for all subjects who took part in the study. Only data from patients who completed at least one arm of the study were retained, and datapoints for only one arm of the study were retained per subject as none completed 4 sessions. Randomisation status is represented by different subplots for each metric (left for Placebo, right for Levetiracetam). Lev = levetiracetam. Proportion correct = proportion of correctly identified items. Identification time = time in seconds identify the correct object. Absolute error = distance between the original item location to the participant’s response location. Localisation Time = the time in seconds to drag the chosen object to its remembered location. More positive values correspond to better performance for proportion correct, and to worse performance for absolute error, identification and localisation time.

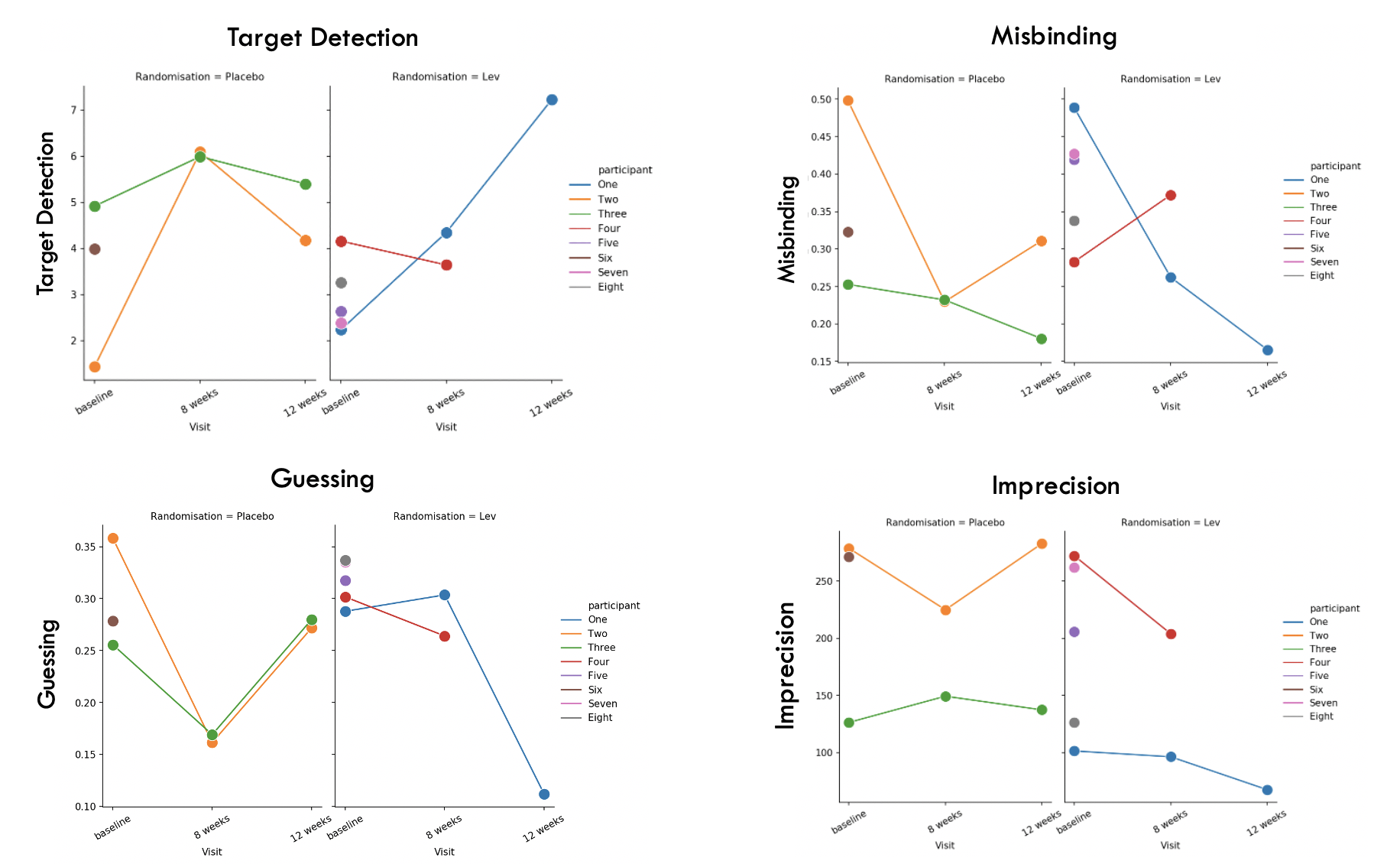

**Supplementary Figure 3 | Performance in the two Fractal condition (mixture model metrics)**

Values are presented as color-coded individual datapoints, for all subjects who took part in the study. Only data from patients who completed at least one arm of the study were retained, and datapoints for only one arm of the study were retained per subject as none completed 4 sessions. Randomisation status is represented by different subplots for each metric (left for Placebo, right for Levetiracetam). Lev = levetiracetam. Target detection = probability of correctly identifying the target. Misbinding = probability of mislocalizing a correctly identified item to the remembered location of another item in the memory array. Guessing = probability of random guessing responses. Imprecision = the width of the distribution of the responses around the target. More positive values correspond to better performance for target detection. More negative values correspond to better performance for misbinding, guessing and imprecision.

**Supplementary Figure 4 | Results of Neuropsychological scales and questionnaires**

No difference was seen across multiple measures between placebo and levetiracetam arms of the study for each participant. LEV = Levetiracetam, NPI = Neuropsychiatric Inventory, BADLS = Bristol Activity of Daily Living Scale, QoL = Quality of life scale (patient and caregiver versions), DSRS = Dementia Severity Rating Scale, EQ5D = Euro-Qol Quality of Life measure (patient, caregiver and proxy versions).

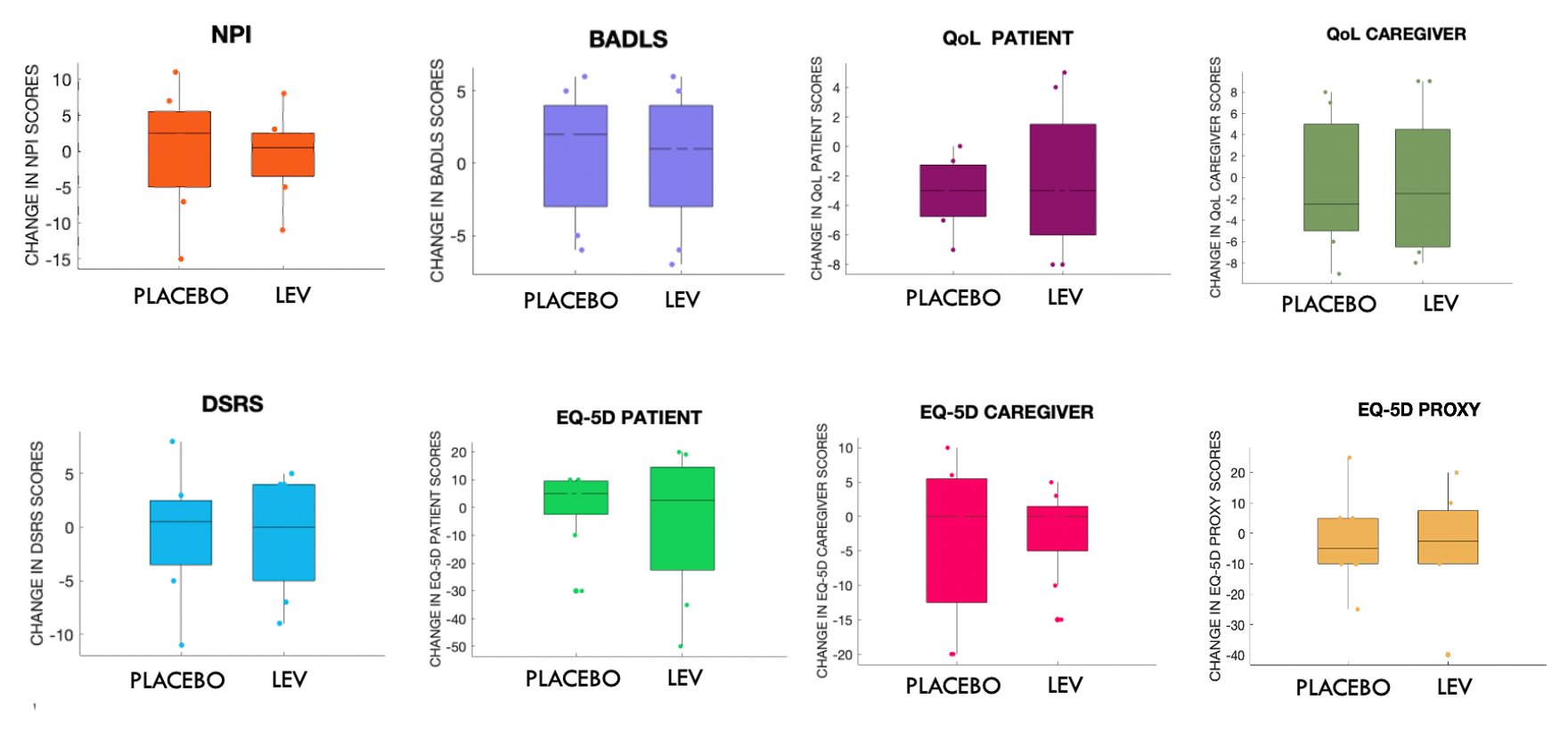

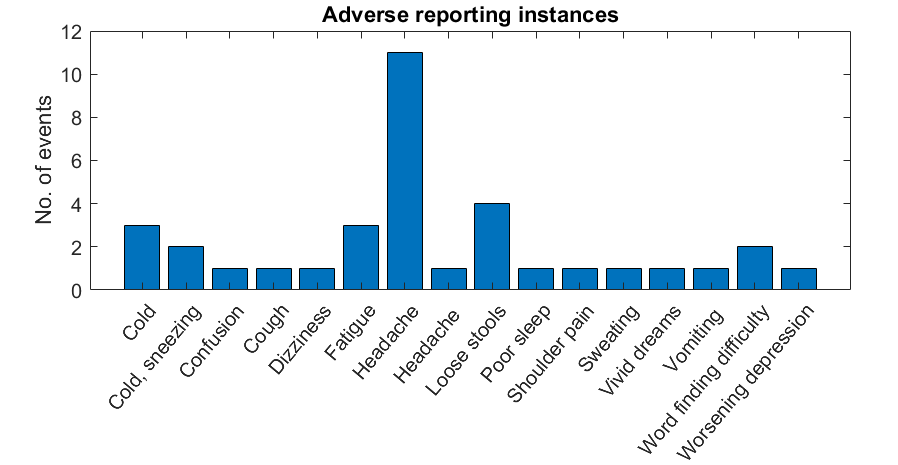

**Supplementary Figure 5 | Adverse reporting instances**

Breakdown of the 35 adverse reporting instances involving a range of non-serious events, the majority of which are recognised as common potential side effects of levetiracetam therapy.

**Supplementary Table 1: Eligibility criteria**

**Inclusion criteria**

**All participants:**

- Participant is willing and able to give informed consent for participation in the trial.

- Participant speaks English as their first language

**Participants with AD:**

- Male or female, 50 years or above.

- Diagnosed with mild to moderate AD (Mini-Mental State Examination score of 10 to 26)

- Meets the National Institute of Aging-Alzheimer’s Association criteria for probable AD (2011)

- Stable dose of current regular medication, including acetylcholinesterase inhibitors if applicable, for at least 4 weeks prior to trial entry.

- Female participants of child bearing potential and male participants whose partner is of child bearing potential must be willing to ensure that they or their partner use effective contraception during the trial and for 3 months thereafter.

- Participant has clinically acceptable blood and urine test results (creatinine clearance > 75 ml/min; liver function tests < 2× upper limit of normal) and ECG that does not demonstrate conduction block or significant ischaemia within 3 months of enrolment.

- In the Investigator’s opinion is able and willing to comply with all trial requirements.

- Willing to allow his or her General Practitioner and consultant, if appropriate, to be notified of participation in the trial.

- Reliable carer willing and available to assist with medication administration as well as to accompany participants during any home visits.

**Carer of participant with AD:**

- Male or female aged 18 and above.

- Principal carer for the participant with AD

- Able to attend all home visits

**Exclusion criteria**

- The participant may not enter the trial if ANY of the following apply.

**Participants with AD**

- Pre-existing diagnosis of epilepsy

- Clinical or laboratory evidence of a cause other than AD as a cause of their dementia

- Laboratory evidence of significant renal impairment (creatinine clearance < 75 ml/minute) or liver dysfunction (liver function tests > 2× upper limit of normal) within the preceding 3 months

- Visual or motor impairment that investigator deems severe enough to impair ability to complete computerised based touchscreen task

- Use of anti-seizure medication for any indication (epilepsy, pain or migraine) within the previous 3 months

- Other severe neurological or medical condition. Examples include significant stroke, heart failure, chronic renal failure, chronic liver failure within last 3 months

- Major depression or other significant behavioural disturbance

- Known allergy to levetiracetam or history of previous adverse reaction to levetiracetam

- Female participant who is pregnant, lactating or planning pregnancy during the course of the trial

- Scheduled elective surgery or other procedures requiring general anaesthesia during the trial.

- Participant with life expectancy of less than 6 months, or is inappropriate for placebo medication.

- Any other significant disease or disorder which, in the opinion of the Investigator, may either put the participants at risk because of participation in the trial, or may influence the result of the trial, or the participant’s ability to participate in the trial

- Participants who have participated in another research trial involving an investigational medicinal product in the past 12 weeks

**Carer of participant with AD**

- Carer has significant medical illness that will preclude adequate data capture during the study

**Supplementary table 2: Potential adverse events**

| Adverse event | Solicited | Unsolicited |
| --- | --- | --- |
| Asthenia |  | ✓ |
| Change in weight |  | ✓ |
| Cough |  | ✓ |
| Effect on mood or behavior | ✓ |  |
| Effect on sleep |  | ✓ |
| Gastrointestinal symptoms |  | ✓ |
| Nasopharyngitis |  | ✓ |
| Rash | ✓ |  |

Levetiracetam is a long-established ASM and potential adverse effects are well known. Common side effects as detailed in the Summary of Product Characteristics (SmPC) were explored with potential participants prior to recruitment. Particular attention was given to whether levetiracetam had an adverse impact on mood or behavior. Table adapted from reference 16.

|  | **Coefficients** | **Standard Error** | **t-value** | **p-value** |
| --- | --- | --- | --- | --- |
| **Fractals 2** |  |  |  |  |
| **Proportion correct** |  |  |  |  |
| **Session** | 0.075 | 0.11 | 0.69 | 0.49 |
| **Drug** | -0.22 | 0.24 | -0.92 | 0.36 |
| **Session * Drug** | 0.1 | 0.15 | 0.65 | 0.52 |
| **Absolute Error** |  |  |  |  |
| **Session** | -64.53 | 43.93 | -1.47 | 0.14 |
| **Drug** | -84.39 | 126.26 | -0.67 | 0.50 |
| **Session * Drug** | 31.41 | 62.13 | 0.50 | 0.61 |
| **Identification time** |  |  |  |  |
| **Session** | 260.85 | 976.63 | 0.27 | 0.79 |
| **Drug** | 6109.7 | 3066.6 | 1.99 | 0.05 |
| **Session * Drug** | -1920.7 | 1381.2 | -1.39 | 0.16 |
| **Guessing** |  |  |  |  |
| **Session** | -0.11 | 0.04 | -2-65 | 0.009 |
| **Drug** | -0.13 | 0.09 | -1.48 | 0.14 |
| **Session * Drug** | 0.11 | 0.06 | 1.87 | 0.06 |
| **Misbinding** |  |  |  |  |
| **Session** | -0.14 | 0.08 | -1.68 | 0.09 |
| **Drug** | -0.13 | 0.19 | -0.69 | 0.49 |
| **Session * Drug** | 0.11 | 0.12 | 0.88 | 0.38 |
| **Imprecision** |  |  |  |  |
| **Session** | 9.90 | 24.30 | 0.41 | 0.68 |
| **Drug** | 18.0 | 92.30 | 0.19 | 0-85 |
| **Session * Drug** | -37.1 | 34.36 | -1.08 | 0.28 |

**Supplementary Table 3 | Fractals 2 Results**

Session = baseline or 8 weeks, Drug = randomization allocation (whether on placebo or levetiracetam), Session x Drug = interaction between session and drug, i.e., whether being on the levetiracetam changed the cognitive outcome compared to placebo at 8 weeks versus baseline. P = p-value, with statistical significance set at p < 0.05.
